# A coding CAG repeat expansion in *EP400* causes Spinocerebellar Ataxia

**DOI:** 10.64898/2026.10.16.26365393

**Authors:** Stijn V. Weckesser, Nataša Dragaševic-Mišković, Mirja Thomsen, Marius Möller, Andona Milovanović, Frauke Hinrichs, Max Borsche, Andreas Dalski, Ana L. de Almeida Marcelino, Sören Franzenburg, Vladimir Kostić, Nikola Kresojević, Shen-Yang Lim, Sebastian Loens, Ana Marjanović, Aleš Maver, Nina Mazalica, Alexander Münchau, Martje G. Pauly, András Salamon, Iva Stanković, Ai Huey Tan, Norbert Brüggemann, Christine Klein, Hauke Busch, Ana Westenberger, Katja Lohmann

**Author notes:** Correspondence to: Katja Lohmann, Institute of Neurogenetics; University of Luebeck; Ratzeburger Allee 160; 23562 Luebeck, Germany. These authors contributed equally to this work (Stijn V. Weckesser, Nataša Dragaševic-Mišković, Mirja Thomsen).

## Abstract

Several polyglutamine-encoding repeat expansions cause spinocerebellar ataxias (SCAs) and other movement disorders. SCAs are genetically heterogeneous with a substantial diagnostic gap.

We used genome sequencing data from 266 individuals of European and Asian ancestry and screened genome-wide for repeat expansions using STRling, a dedicated repeat expansion caller. The cohort comprised patients with movement disorders, predominantly dystonia and ataxia, as well as unaffected participants. Further, we screened another 436 individuals with ataxia and 427 ethnically matched controls through fragment analysis of the most promising candidate locus in *EP400*. We applied targeted long-read sequencing to expansion carriers and one control individual to resolve the sequence structure of the repeat locus. Finally, we used microsatellite markers in chromosome 12q24.32–33 to test for a common founder.

A genome-wide outlier analysis in STRling revealed a polyglutamine-encoding repeat expansion (GRCh38 chr12:132,062,525–132,062,590) in the E1A binding protein p400 (*EP400*) gene in two patients with ataxia. Through screening of additional ataxia patients, we identified in total 18 patients from 14 families with heterozygous repeat expansions in *EP400.* All expansion carriers were from Serbia. The repeat number in affected individuals ranged from 37 to 55, whereas the longest control allele had 32 repeats. We demonstrated that non-expanded alleles carried a CAA interruption within the 5′ CAG tract – this interruption was lost in expanded alleles. Expansion carriers have pure SCA with onset mainly in their 40s and 50s. There is an inverse correlation between the number of repeats and age at onset. The *EP400-*encoded protein plays an essential role in chromatin remodeling, epigenetic regulation and DNA repair. Pathogenic sequence variants in *EP400* have previously been implicated in a neurodevelopmental disorder.

Our study identifies repeat expansions in *EP400* as a novel and frequent cause of ataxia in Serbian patients. This finding further increases the diagnostic yield of ataxia testing and warrants investigation in ataxia patients worldwide. We add *EP400* to the growing list of ataxia-associated repeat loci, which warrants inclusion in diagnostic ataxia panels.

## Introduction

Short tandem repeats (STRs) are stretches of DNA sequence motifs of two to six base pairs repeated multiple times in succession.^1^ STRs can undergo expansion, i.e., an increase in the number of repeat-motif copies, giving rise to a group of predominantly neurological conditions collectively referred to as repeat expansion (RE) disorders.^1,2^ The genomic location of an expanded STR, its motif sequence and the length of the expanded allele determine its pathogenicity. A large proportion of RE disorders manifest as movement disorders. For instance, coding CAG expansions, which translate into elongated polyglutamine tracts, are an established cause of several hereditary ataxias and of Huntington’s disease.^1,3^ As of September 2026, the STRipy database lists 61 RE disorders, of which 17 are associated with ataxia and 11 linked to a CAG RE.^4,5^

Adult-onset cerebellar ataxias, including spinocerebellar ataxias (SCAs), are a clinically and genetically heterogeneous group of progressive neurodegenerative disorders.^6,7^ Affected individuals typically present with gait ataxia, ocular motor abnormalities and dysarthria, often accompanied by non-cerebellar features.^6^ More recently, advances in sequencing technology and bioinformatic analysis have enabled the discovery of further, often deep intronic, REs, including those in *FGF14* and *RFC1*, in a considerable number of patients with ataxias.^2,8^ However, a substantial proportion of affected individuals remains without a molecular diagnosis, with a reported median diagnostic yield of about 50% for hereditary ataxias.^9^ The relevance of REs to other movement disorders is less pronounced. There are a few reports linking REs in *FGF14*, *RFC1* and *NOTCH2NLC* to parkinsonism.^10–12^ Further, intermediate-length CAG expansions in *ATXN2* are recurrently found in families with Parkinson’s disease. CAG repeat expansions in *HTT* lead to Huntington’s disease (HD), which is characterized by chorea, dystonia and cognitive decline.^13^ No RE has been established as a cause of isolated dystonia.^1^

In order to further elucidate the role of REs in movement disorder patients, we used short-read genome sequencing data from 266 individuals and applied STRling, a catalogue-free tool for genome-wide detection of repeat expansions (Fig. 1). Fragment length analysis and targeted Nanopore amplicon sequencing of the top candidate across 14 families of Serbian origin resolved the repeat length, internal structure and a shared haplotype. Our findings implicate *EP400* as a novel repeat expansion locus in ataxia.

**Figure 1.**
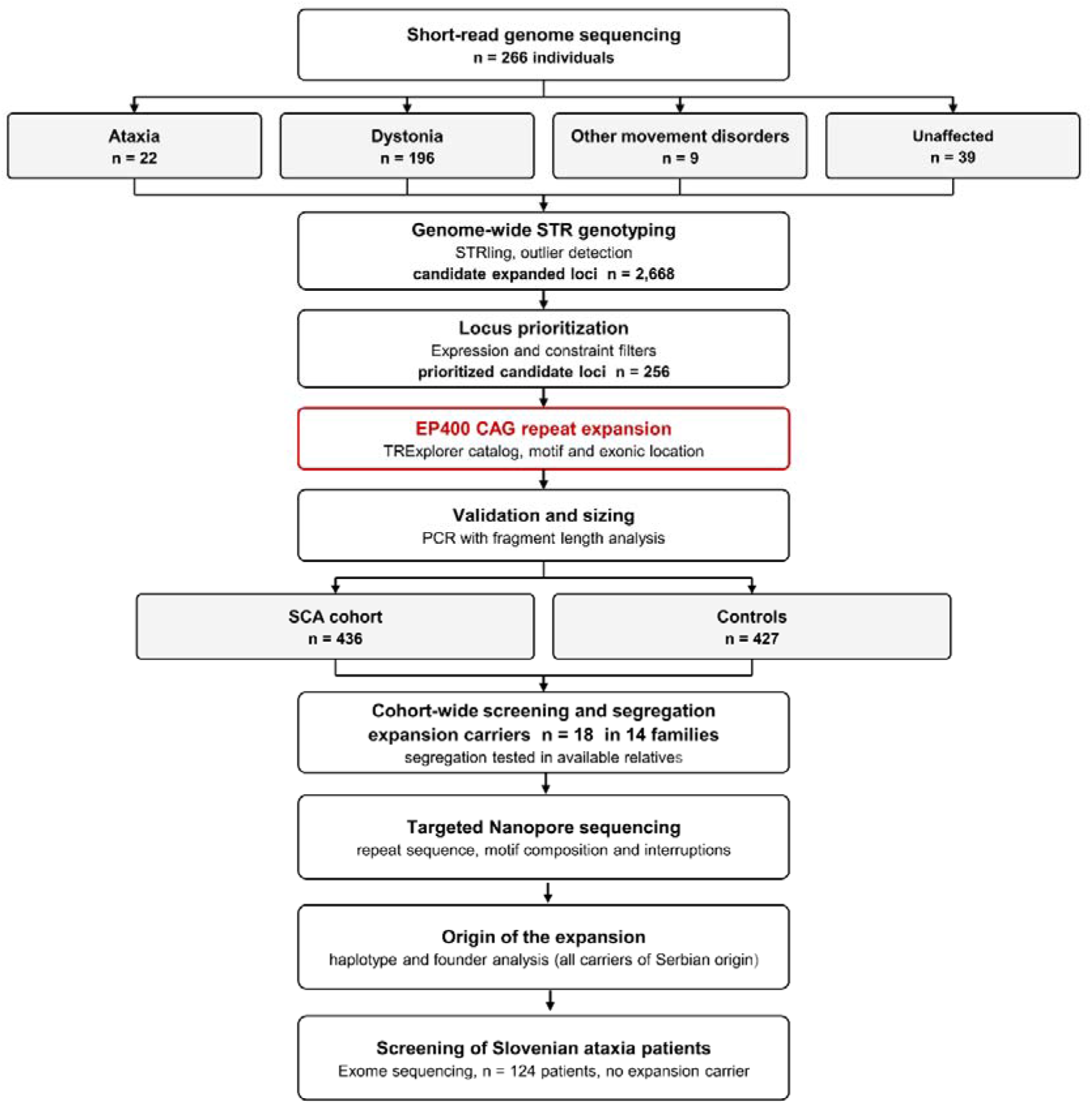
Workflow of the identification and characterization of the *EP400* repeat expansion. The different steps carried out in this study are illustrated.

## Material and Methods

### Discovery cohort and short-read genome sequencing

For the discovery cohort, we used short-read genome sequencing data from 266 individuals, generated on an Illumina NovaSeq platform, as recently described.^14^ All patients were examined by experienced neurologists. Dystonia was the presenting phenotype in 73.7% of genome-sequenced participants, ataxia in 8.3% and other neurological disorders in 3.4%. The remaining 14.6% were unaffected individuals, who either contributed to segregation analysis within families or served as healthy controls. Among all participants, 50.3% were female. Across index patients with dystonia as the leading phenotype, 47.8% were of European and 52.2% of Asian ancestry. Family history was positive in 29.3% of them, and the median age at onset was 12 years (interquartile range [IQR]: 6–28 years, range: 0–78 years). Among index patients with ataxia as the leading phenotype, 68.4% were of European and 31.6% of Asian ancestry. Family history was positive in 31.6% of them, and median age at onset was 35 years (IQR: 32–43 years, range: 3–50 years). Median age at examination was 50 years in affected individuals (IQR: 33–60 years, range: 16–80 years) and 55 years in unaffected individuals (IQR: 55–57 years, range: 53–60 years).

Ethical approval was obtained from the local ethics committee or institutional review board, and all procedures followed the ethical standards of the relevant institutional and national research committees. All participants gave written informed consent.

### STRling and filtering

To systematically screen the cohort for potentially pathogenic REs, we analyzed the genome sequencing data using STRling.^15^ After generating an STR index from the reference FASTA file, informative read pairs were extracted from each sample and combined across the cohort to identify STR loci that could subsequently be genotyped in each individual. We then used STRling’s outlier analysis to highlight expanded alleles relative to the remainder of the cohort. We retained only STRling calls whose *P*-value, adjusted for multiple testing within each sample using the Benjamini– Hochberg method, was below 9.9 × 10 .^15,16^ This yielded 2,668 different candidate STR loci across all individuals. We then excluded loci meeting any of the following eight criteria: (i) low read depth (< 10 reads); (ii) location on alternative contigs; (iii) repeat unit length < 3 bp; (iv) intergenic location; (v) location in a gene without detectable expression in brain tissue; (vi) *sum_str_log* < 1.0, defined as the log of the depth normalized sum of STR repeat units across all reads assigned to the locus; (vii) detection in a single individual only; and (viii) previously reported pathogenic STRs in STRipy. In addition, loci whose estimated allele length (i.e., repeat number) did not exceed that of any unaffected individual by more than one repeat unit were considered unlikely to be pathogenic. For each of the 256 loci that passed these filters, we compared the estimated allele size in repeat units with the allele sizes reported for the corresponding locus in TRExplorer.^17^ Where available, we used short-read allele sizes from TRExplorer for this comparison. We prioritized STRs in coding regions and STRs carrying a known pathogenic motif, as both features are associated with an increased odds ratio for pathogenicity.^18^ The strongest candidate to emerge from this was a coding CAG repeat in exon 47 of *EP400* (NM_015409.5), for which STRling estimated allele sizes in two of our patients exceeding those reported in TRExplorer.^17^ We then inspected this locus visually in genome data (Fig. 2 A and B) using the Integrative Genomics Viewer (IGV; v2.19.7) to identify soft-clipped reads composed of CAG repeats adjacent to the polyglutamine encoding region (GRCh38 chr12:132,062,525–132,062,611 with the reference sequence of (CAG)_6_(CAA)_2_(CAG)_14_(CAA)_1_(CAG)_4_(CAA)_1_(CAG)_1_).^19^

**Figure 2.**
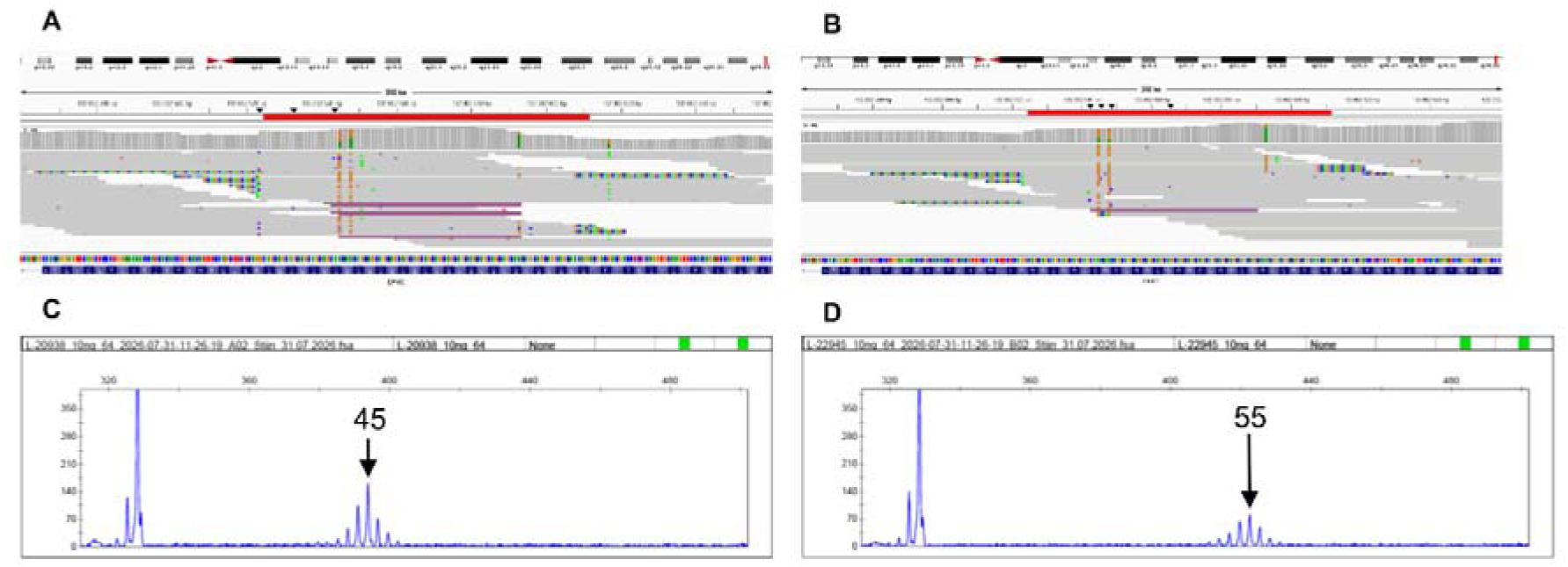
Visualization of the *EP400* repeat expansion in genome sequencing data and fragment length analysis of *EP400* alleles. (**A** and **B**) Integrative Genomics Viewer (IGV) screenshots of the *EP400* repeat in Individuals L-20938 (**A**) and L-22945 (**B**) from the discovery cohort. Short-read genome sequencing data from these individuals revealed soft-clipped reads flanking the polyglutamine-encoding region. The red bar at the top marks the complete polyglutamine-encoding region in exon 47 of *EP400* (GRCh38 chr12:132,062,525–132,062,611). (**C** and **D**) The electropherograms of Individuals L-20938 (**C**) and L-22945 (**D**) show the typical pattern of heterozygous expansion carriers. There is one smaller, rather single peak for the interrupted, non-expanded allele of 23 repeats and a larger peak for the expanded allele that indicates instability, with less pronounced peaks at ±1 and ±2 repeats (45 repeats in **C** and 55 in **D**). Repeat number and sequence were confirmed by sequencing. The x-axis shows the fragment length in base pairs and the y-axis the relative fluorescence intensity.

### Validation cohort screening by PCR and fragment analysis

Following the detection of expanded *EP400* alleles in the discovery cohort, we screened an independent in-house sample of 436 individuals with ataxia (43.4% female; 55.8 % of German ancestry, 43.3 % of Serbian, 0.9 % of Hungarian ancestry) and 427 control individuals (50.6 % female; 72.4 % German, 27.6 % Serbian). German controls were recruited within the EPIPARK study, and Serbian control individuals through Serbian neurologists.^20^ All patients and control individuals provided written informed consent prior to inclusion. For *EP400* repeat screening of these additional ataxia patients and controls, we amplified the repeat-containing (GRCh38 chr12:132,062,525–132,062,611) and adjacent regions by PCR with a fluorescently labelled forward primer and Taq DNA polymerase (MP Biomedicals), followed by fragment length analysis (Fig. 2 C and D) on a 3500XL Genetic Analyzer (Applied Biosystems) using POP-7 polymer and GeneScan 600 LIZ as size standard. Alleles were called and scored in GeneMapper v6.1 (Applied Biosystems). Fragment lengths were converted into repeat units, focusing on the first repeat stretch from (GRCh38 chr12:132,062,525–132,062,590, i.e., (CAG)_6_(CAA)_2_(CAG)_14_) in analogy to the analysis in *ATXN1*, *ATXN2* and *HTT*. We subtracted the other portion of the amplicon from the total fragment length and then divided the resulting length by 3, corresponding to the repeat motif length, to obtain the repeat number. Upon long-read sequencing (see below) and Sanger sequencing of four homozygous samples, we corrected the repeat number (+3). This was necessary since the fragment analysis underestimated allele sizes by three repeat units compared with sequence-based sizing, consistent with the systematic underestimation previously observed for the CAG repeat in *ATXN1*.^21^ Primer sequences and detailed PCR conditions are provided in the Supplementary material.

### Sanger sequencing and targeted Nanopore amplicon sequencing

To characterize the sequence composition of the top candidate, we performed Sanger sequencing and long-read sequencing. For Sanger sequencing, we used the same primers as for fragment analysis, and the Phusion Flash High-Fidelity PCR Master Mix (Thermo Fisher Scientific) was applied to four samples with homozygous repeat length to exactly determine the repeat number. For long-read sequencing, the repeat-containing region was amplified from DNA samples of 17 patients (for one sample with an expanded allele, DNA amount was not sufficient for long-read sequencing) and one control sample. For this, we used the Phusion Flash High-Fidelity PCR Master Mix (Thermo Fisher Scientific) and the same locus-specific primers as used for the fragment analysis (without fluorescent label; see Supplementary material). Amplicons were purified using AMPure XP beads (Beckman Coulter). For each sample, 200fmol of purified amplicon DNA was used for library preparation following the Oxford Nanopore Technologies (ONT) protocol SQK-NBD114.24. Samples were multiplexed using the Native Barcoding Kit 24 V14 (ONT) and the library was sequenced on a MinION device using an R10.4.1 flow cell (∼3h, super-accurate base calling, Q-score ≥10). Nanopore reads were processed using an in-house Nextflow-based pipeline. Reads were quality-filtered using NanoFilt and adapter-trimmed with Porechop, followed by alignment to the GRCh38 human reference genome using minimap2. Alignments were sorted and indexed using SAMtools. Repeat-spanning reads were clustered into alleles and allele-specific consensus sequences were generated using STRdust in unphased mode, using up to 10,000 reads for allele clustering and 500 reads for consensus generation.^22^ Median read length was used to determine repeat number. Repeat motif composition and interruptions were visualized using aSTRonaut, and reads were additionally visually inspected in IGV.^18,22^

### Haplotype analysis

We genotyped 11 STR markers flanking the repeat (10 established ones [D12S1639, D12S1714, D12S367, D12S97, D12S63, D12S392, D12S1723, D12S1628, D12S357 and D12S1638], and one new marker located 0.3Mb downstream of the repeat towards the end of the chromosome) using fragment length analysis. Primer sequences of the new marker are provided in the Supplementary material.

### Targeted *EP400* genotyping by ExpansionHunter in Slovenian patients

To further explore the role of RE in *EP400* in Slovenia, a Southeast European country, we analyzed diagnostic exome sequencing data from the University Medical Center in Ljubljana, the reference center for movement disorders in Slovenia. Using ExpansionHunter, we genotyped the *EP400* repeat locus in 124 unrelated adults with ataxia recruited in Slovenia (64 male and 60 female; median age at onset 52 years, range 17 to 85 years).^23^ All genotype calls passed quality control, with a median read depth of 84.6× at the locus. Genome data from this cohort had been generated previously under broad consent for research use.

### Statistics

Statistical analyses were performed in R (version 4.5.2), and figures were generated with ggplot2. We summarized continuous variables as medians with interquartile ranges and categorical variables as counts and percentages. We compared allele length distributions between patients and controls in the validation cohort using a two-sided Mann–Whitney U-test. The relationship between age at onset and repeat length was assessed by simple linear regression, with age at onset as the dependent variable and the length of the expanded allele as the predictor, using the lm function in R.

## Results

### STRling analysis

Genome-wide profiling with STRling yielded 256 candidate loci that passed all filtering criteria (Supplementary Table S1).^15^ Only one candidate met both prioritization features: A CAG repeat in exon 47 of *EP400* (NM_015409.5), which lies within a coding exon and consists of a motif (CAG) known to be pathogenic at other loci. STRling called an expanded allele at this locus in two unrelated individuals, both of whom presented with SCA and were from Serbia. Inspection of the aligned reads showed soft-clipped ends composed entirely of CAG units immediately adjacent to the polyglutamine-encoding tract (Fig. 2 A and B), providing read-level support for an expansion in both individuals.

### Cohort screening of ataxia patients for repeat expansions in ***EP400***

Screening of independent in-house ataxia samples of German and Serbian ancestry by fragment length analysis identified an expanded *EP400* allele in another 16 of 436 patients with ataxia (3.7%), but in none of 427 ethnically matched control individuals. Together with the two index patients from the genome-sequenced patient group, 18 patients from 14 families with SCA, all of Serbian ancestry, were found. Expanded alleles in affected individuals ranged from 37 to 55 pure CAG repeat units. In controls, the longest allele comprised 32 repeat units. Alleles of 33 to 36 repeat units were observed in neither patients nor controls, and currently define the “gray zone”. The distribution of allele lengths in both groups is shown in Fig. 3. Allele lengths differed significantly between individuals with ataxia and controls (Mann–Whitney U-test, *U* = 352952.5, *P* = 0.01498; R 4.5.2).

**Figure 3.**
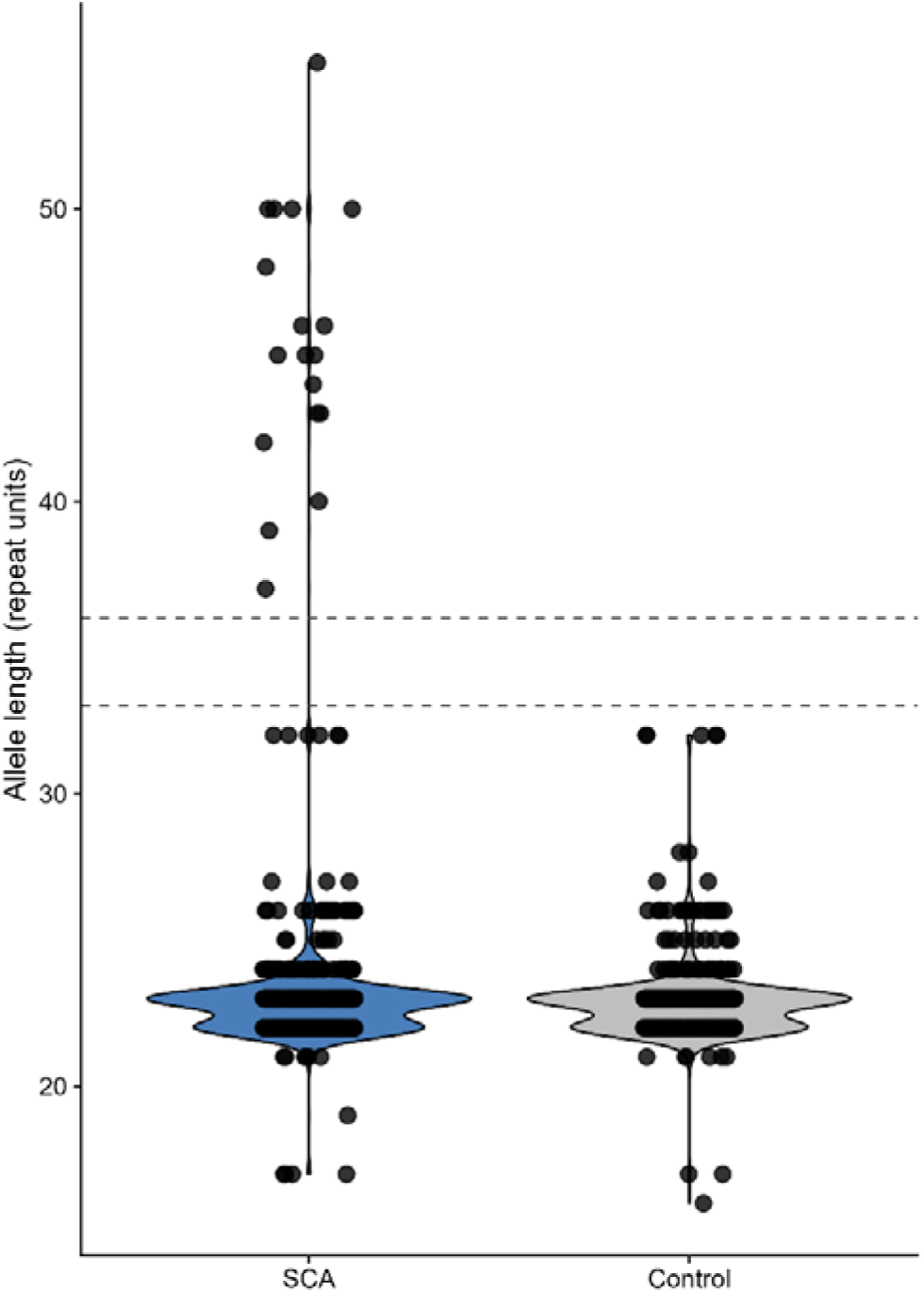
Distribution of *EP400* repeat lengths in ataxia patients and in control individuals. Violin plots showing the allele length distribution in 436 individuals with ataxia and 427 controls, with individual alleles overlaid as points. Median allele length was 23 repeat units in both groups (IQR 22 to 23). The lower dashed line marks the upper limit of the range in which alleles were present in both groups. The upper dashed line marks the lower limit of the range in which expanded alleles were detected in affected individuals. Alleles falling in the “gray zone” between the two lines (33–36 repeat units) were observed in neither group, and it therefore remains unclear whether alleles of this size are pathogenic or benign.

Next, we explored diagnostic exome sequencing data from 124 Slovenian ataxia patients for RE in *EP400* using ExpansionHunter.^23^ The longest allele per patient ranged from 22 to 28 repeats; thus, none of the Slovenian patients carried an RE in *EP400*.

Most of the newly identified *EP400* expansion carriers (n=15) had previously been tested for REs in *ATXN1, ATXN2, ATNX3, ATXN7*, *CACNA1A*, *FGF14* and *RFC1*.^24^ In these patients and in the two genome-sequenced carriers, other genetic causes of ataxia had been excluded by comprehensive analyses. Our sample included eleven multiplex families; for three of these families, DNA from 2–3 affected individuals was available for testing (Fig. 4). Within families, all affected individuals who were available for testing (all from the same generation) also carried the expansion at a size of +/- 1 repeat unit. The observed segregation pattern is consistent with autosomal dominant inheritance and high penetrance. In the Serbian cohort, this expansion accounts for about 6.5% of patients, much higher than in other polyglutamine expansion-containing SCA genes. When looking at Serbian patients with a positive family history compatible with autosomal dominant inheritance, the diagnostic yield increases further to almost 20% (n=14/74).

**Figure 4.**
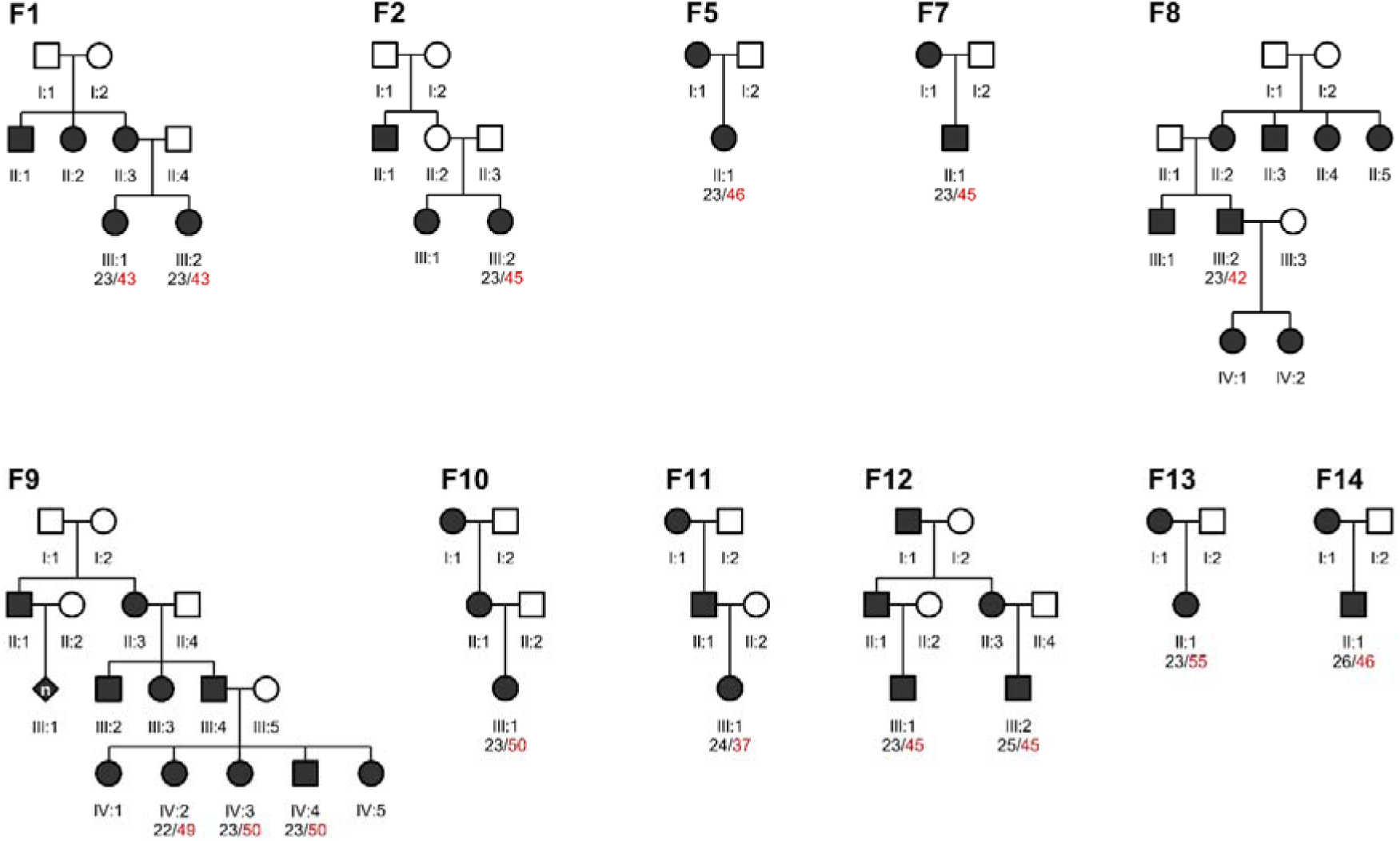
Pedigrees of the 11 multiplex families with affected members harboring an *EP400* repeat expansion. Squares denote males, circles females and diamonds family members of unknown sex. Filled symbols indicate individuals reported or examined as affected; open symbols indicate unaffected individuals. For all genotyped individuals, the repeat number for both *EP400* alleles is given below the individual pedigree number; the expanded allele is shown in red. Relatives reported as affected by family history were not available for genetic testing. “n” inside a symbol indicates that the number of individuals represented is unknown. Pedigrees are not shown for the three expansion carriers with a negative family history.

### Genotype–phenotype correlation

From a clinical perspective, *EP400* RE carriers had a mean age at onset of 48 years (IQR: 42 to 56 years, range: 14 to 61 years) and all showed limb and gait ataxia, cerebellar dysarthria and pyramidal signs. Ocular motor abnormalities such as gaze-evoked nystagmus (83.3%) and dysmetric saccades (39.9%) were present in many patients. RE in *EP400* are clinically characterized by an adult-onset cerebellar syndrome with facultative upper motor neuron signs and cerebellar atrophy. Available clinical data of all individuals carrying an expanded allele are summarized in Table 1.

**Table 1.** Demographic, clinical and genetic findings in 18 patients from 14 families carrying an *EP400* repeat expansion.

| Family ID | ID | *EP400* repeat number<sup>b</sup> | Family history | Disease duration (years) | Upper limb ataxia | Lower limb ataxia | Gait ataxia | Cerebellar dysarthria | Dysphagia | Walking assistance | Spasticity | Increased deep tendon reflexes | Gaze-evoked nystagmus | Dysmetric saccades | SARA score | Brain MRI |
| --- | --- | --- | --- | --- | --- | --- | --- | --- | --- | --- | --- | --- | --- | --- | --- | --- |
| F1 | P1 | 23/43 | pos | 20 | + | + | + | + | + | – | + | + | + | + | 21 | Cerebellar atrophy |
| F1 | P2 | 23/43 | pos | 13 | + | + | + | + | + | – | – | + | + | + | 16 | Cerebellar atrophy |
| F2 | P3 | 23/45 | pos | 13 | + | + | + | + | – | + | – | + | + | – | ND | ND |
| F3 | P4 | 22/44 | neg | 15 | + | + | + | + | – | – | + | + | + | – | 21 | Global brain atrophy |
| F4 | P5 | 24/40 | neg | 5 | + | + | + | + | – | – | + | + | ND | ND | ND | Global brain atrophy |
| F5 | P6 | 23/46 | pos | 10 | + | + | + | + | – | + | + | + | + | + | 23 | Cerebellar atrophy |
| F6 | P7 | 24/50 | neg | 8 | + | + | + | + | – | – | + | + | + | – | 15 | Cerebellar atrophy |
| F7 | P8 | 23/45 | pos | 8 | + | + | + | + | – | – | + | + | + | – | 8 | Global brain atrophy |
| F8 | P9 | 23/42 | pos | 10 | + | + | + | + | + | + | + | + | + | + | 20 | Unremarkable |
| F9 | P10 | 22/49 | pos | 5 | + | + | + | + | – | – | + | + | + | – | ND | Unremarkable |
| F9 | P11 | 23/50 | pos | 34 | + | + | + | + | – | – | + | + | + | – | ND | Global brain atrophy |
| F9 | P12 | 23/50 | pos | 10 | + | + | + | + | + | – | + | + | – | – | ND | Cerebellar atrophy |
| F10 | P13 | 23/50 | pos | 18 | + | + | + | + | – | + | + | + | + | – | ND | Global brain atrophy |
| F11 | P14 | 24/37 | pos | 4 | + | + | + | + | + | – | + | + | – | + | 8 | Global brain atrophy |
| F12 | P15 | 23/45 | pos | 16 | + | + | + | + | – | – | + | + | + | – | 22 | Global brain atrophy |
| F12 | P16 | 25/45 | pos | 4 | + | + | + | + | + | – | + | + | – | + | 13 | Global brain atrophy |
| F13 | P17 | 23/55 | pos | 10 | + | + | + | + | – | – | + | + | + | + | 13 | Cerebellar atrophy |
| F14 | P18 | 26/46 | pos | 3 | + | + | + | + | – | – | + | + | + | – | ND | Cerebellar atrophy |

All patients were of Serbian ancestry. F = female; ID = identifier; M = male; ND = no data available; neg = negative family history; pos = positive family history; SARA = Scale for the Assessment and Rating of Ataxia; + = present; – = absent.
<sup>a</sup>Scored on clinical ocular motor examination.
<sup>b</sup>Repeat sizes are given as shorter/longer allele in repeat units.
<sup>c</sup>Age at which the first motor symptom was reported by the individual or a first-degree relative.

Regarding the variable age at onset, an inverse relationship between repeat length and age at onset is a recurring feature of RE disorders, especially for CAG repeats.^1,2^ To assess whether the *EP400* repeat behaves accordingly, we performed a linear regression, with age at onset as the dependent variable and the length of the longer allele as the independent variable. Longer alleles were associated with an earlier age at onset (R² = 0.46, *P* = 0.002, slope −1.965; Fig. 5). Within the analyzed range of 37 to 55 repeat units, each additional repeat unit corresponded to an earlier onset by approximately two years.

**Figure 5.**
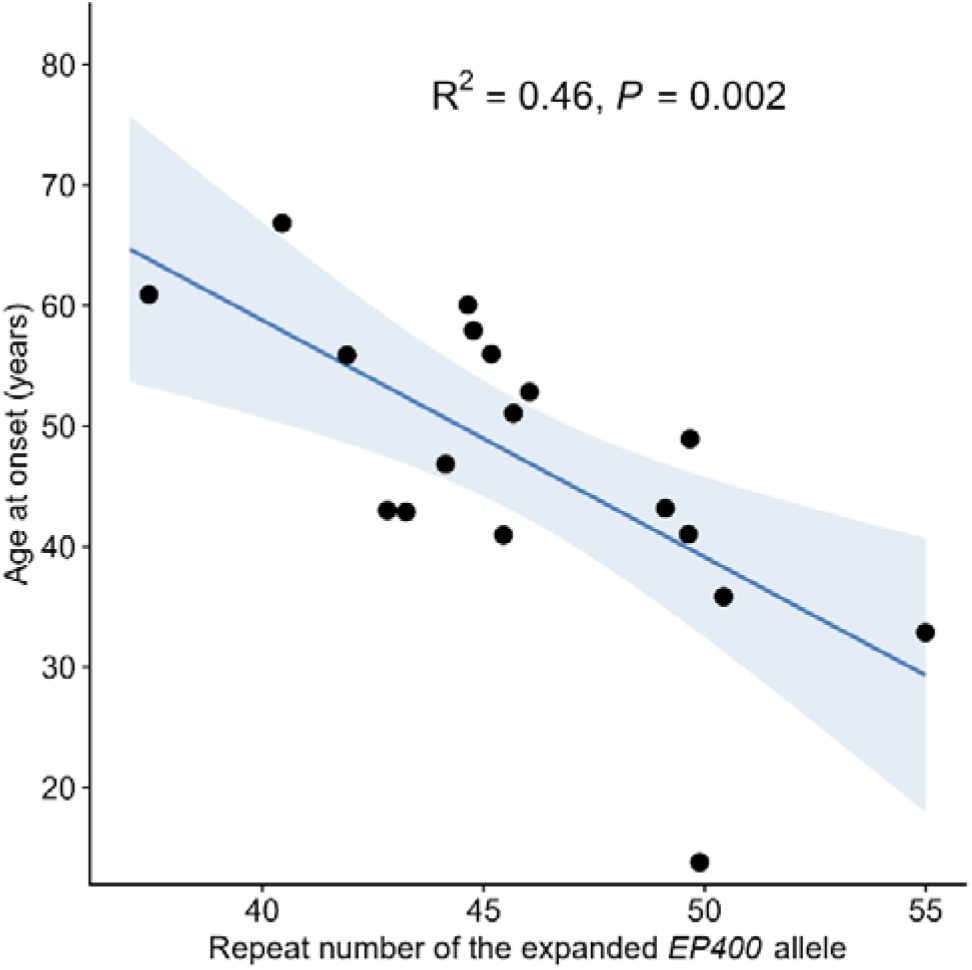
Age at onset correlates inversely with expanded *EP400* repeat length. Age at onset plotted against the length of the expanded *EP400* repeat in *n* = 18 carriers. Each dot represents one individual; the line shows the linear regression with its 95% confidence interval. Repeat length explained a substantial part of the variance in age at onset (R² = 0.46, *P* = 0.002), with each additional repeat unit corresponding to an onset 1.965 years earlier.

#### Motif structure of expanded and non-expanded *EP400* alleles

Sanger sequencing and long-read sequencing resolved the internal structure of the repeat. The most common, non-expanded allele comprised a total of 30 repeat units with the structure (CAG)_6_(CAA)_2_(CAG)_15_(CAA)_1_(CAG)_4_(CAA)_1_(CAG)_1_, highly similar to the one reported in the reference genome. Long-read sequencing showed that all expanded alleles had lost the (CAA)_2_ block in the 5′ region of the repeat, while retaining the two single CAA interruptions at the 3′ end. This indicates that the internal, uninterrupted sequence of the tract, and not its length alone, should be taken into account when this locus is genotyped. There was some variability in the exact position and number (n=1–2) of the 5’-region interruption (Fig. 6). This is similarly observed in other polyglutamine disorders, in which the internal sequence composition, i.e., the presence of interruptions, of the repeat tract determines its stability and modifies its pathogenicity.^21,25–27^ The comparison of *EP400* RE with other CAG REs in *ATXN1* (SCA1), *ATXN2* (SCA2) and *HTT* (HD) is shown in Fig. 6. As in *EP400*-related ataxia, the expanded alleles of SCA1 and SCA2 patients have lost their CAA interruptions in the 5’ regions. In contrast, the CAA repeats in the 3’ region of the repeat do not prevent the expansion and are also present in the expanded CAG repeat in *HTT*.

**Figure 6.**
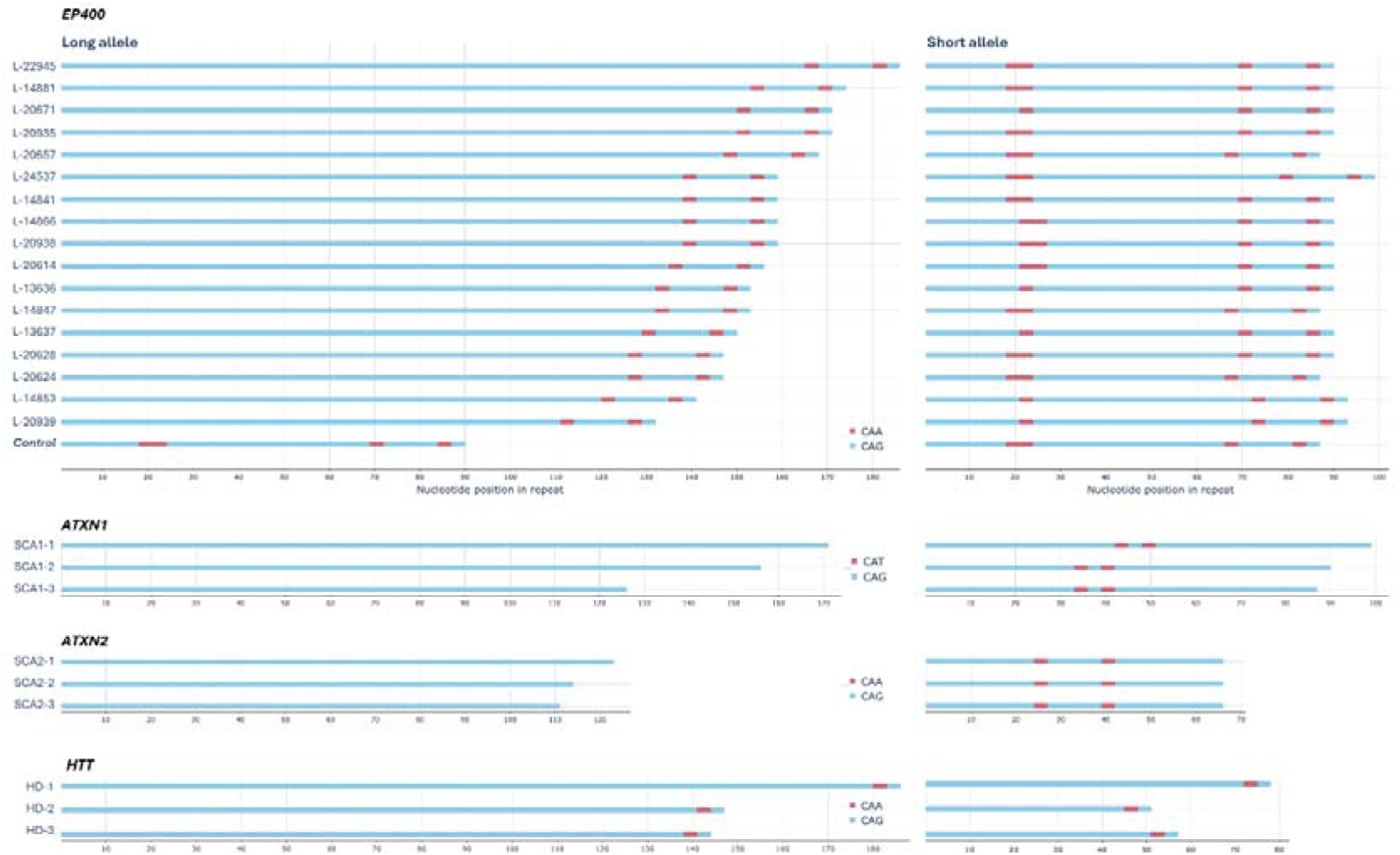
Repeat motif composition at the *EP400* repeat locus compared with established CAG repeat expansion disorders. Repeat structures were determined by Oxford Nanopore long-read sequencing. For each individual – the 17 *EP400* expansion carriers reported here for whom DNA was available, a healthy Serbia control without an *EP400* repeat expansion, and three in-house patients each with a pathogenic repeat expansion in *ATXN1* (SCA1), *ATXN2* (SCA2) or *HTT* (HD), respectively, used for comparison – the longer allele is shown on the left and the shorter allele on the right. Long-read sequencing of all *EP400* expansion carriers revealed loss of the 5′ CAA interruption in the expanded alleles, resulting in a long uninterrupted CAG tract, while the interruption motifs at the 3′ end were retained. A similar pattern is observed in expanded *ATXN1* and *ATXN2* alleles, whereas the 3′ CAA interruption in expanded *HTT* alleles is retained, as were the 3′ interruption motifs in *EP400*.

For counting of the repeat size, we therefore focused on the 5’ region of the repeat, which corresponds in non-expanded alleles usually to (CAG)_6_(CAA)_1-2_(CAG)_14-15_ and in expanded alleles to (CAG)_>37_.

### Haplotype analysis

Notably, all individuals carrying an expanded allele were of Serbian ancestry; none was found among the tested patients from Germany, Hungary, Malaysia or Slovenia. This suggests a common Serbian founder. We therefore genotyped 11 STR markers flanking the repeat. Allele identity was highest for the markers closest to the repeat and declined with increasing distance. Specifically, the three nearest markers (D12S1723, D12S1628 and a custom marker) were identical in all carriers, defining a common haplotype across 0.8 Mb, whereas the most distant marker was identical in only 9 of 18 carriers (50%). The same haplotype was shared across all 11 markers by 9 of 18 individuals (50%) and across a 3.0 Mb interval (2.7 Mb 5′ and 0.3 Mb 3′ of the repeat) by 15 of 18 expansion carriers (83.3%). Haplotype data together with the presumed founder haplotype are shown in Fig 7. This pattern is consistent with inheritance from a common founder.

**Figure 7.**
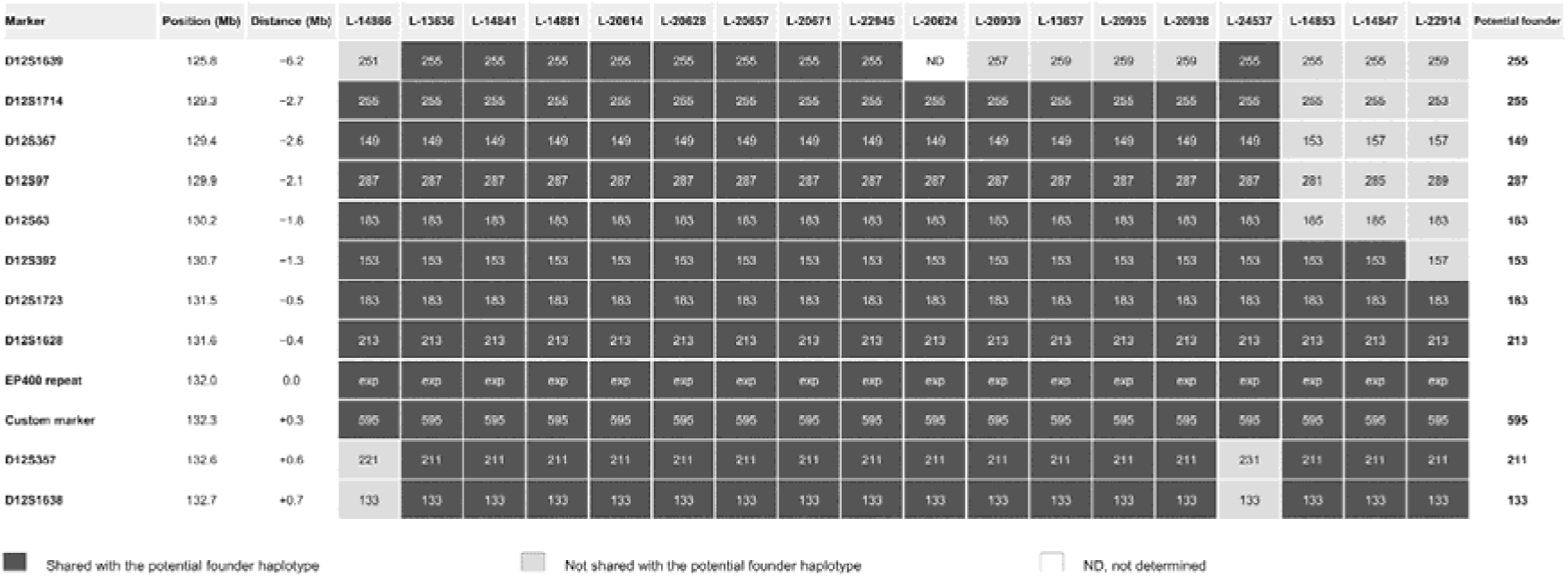
Haplotype analysis of STR markers flanking the EP400 repeat expansion. Haplotypes of 18 individuals carrying the *EP400* repeat expansion at 11 microsatellite markers flanking the repeat. Markers are ordered by chromosomal position on chromosome 12q24.32–33. The potential founder allele is defined as the most frequent allele per marker among expansion carriers and shared among siblings. Individuals are ordered by the extent of their deviation from this founder haplotype, with carriers of smaller alleles shown on the left and carriers of larger alleles on the right. Dark grey marks the segment shared with the potential founder haplotype. Light grey marks all deviating markers, indicating that the region is no longer shared with the founder haplotype. The *EP400* repeat is also indicated. Allele sizes are given in base pairs. Positions refer to GRCh38, and distances are given relative to the *EP400* repeat (negative, centromeric; positive, telomeric). ND, not determined

## Discussion

In this study, we identified a CAG repeat expansion in *EP400* as a novel cause of SCA. Interestingly, a recent population-scale analysis of STR variability in long-read genomes proposed *EP400* as a candidate locus for a potentially pathogenic RE, based on its Pure Length Variability Index, a measure of allelic variability at a given locus, combined with the detection of individual allele-length outliers in two index patients.^28^ According to our data, RE in *EP400* are frequent among Serbian ataxia patients, but rare among other European patients, including other Southeast Europeans and Germans. Accordingly, STR-based haplotype analysis indicated a common founder.

Given the common founder haplotype shared by most carriers, the observed range of *EP400* repeat lengths (37–55 repeat units) may be caused by intergenerational repeat instability. Due to a lack of respective patient samples, i.e., parent–offspring pairs, we were not able to investigate the intergenerational stability of this RE. Among the sibling and cousin pairs, we found a maximum allele difference of one repeat. This is consistent with the findings in other trinucleotide repeats, which behave as dynamic mutations and tend to expand further upon transmission once a gene-specific threshold is exceeded.^2^ Additional expansion carriers and further fine-mapping of the shared region will elucidate the age of this potential founder. Long-read sequencing resolved the internal structure of the repeat and showed that all expanded alleles lacked the 5′ CAA interruption present in non-expanded alleles. Interruptions of the repeat tract play an important role in several RE disorders, including CAG but also GGC repeats.^7,29^ For example, *in vitro*, uninterrupted (CAG)_n_ tracts form stable non-B DNA structures, including hairpins, triplexes, quadruplexes and slipped-strand structures, which can interfere with DNA replication, repair and recombination.^30^ In *ATXN2*, it has been shown that CAA interruptions confer stability on the repeat, while, in contrast, pure tracts are more unstable and more prone to expansion.^31^ Further, in *Drosophila* models of *ATXN3*, CAA interruptions have been proposed to reduce toxicity at the RNA level, indicating that the interruption pattern itself, and not only overall repeat length, modulates pathogenicity.^32,33^ We therefore speculate that the loss of the first CAA island in *EP400* contributes to reduced stability and increased toxicity of the repeat at this locus, as well.

This complex repeat structure also has consequences for how the locus is genotyped. The population-scale analysis only covers the longest pure CAG repeat (GRCh38 chr12:132,062,548–132,062,611), whereas our data indicate that the variability occurs in the 5’ polyglutamine-encoding region (GRCh38 chr12:132,062,525–132,062,590).^28^ Therefore, this 5’ region needs to be assessed, since the loss of the 5′ CAA interruption may itself contribute to pathogenicity. Our findings underscore the importance of analyzing the length and interruption pattern when assessing an RE.

*EP400* encodes a p400 protein that acts as an ATP-dependent chromatin remodeling scaffold and is a core component of the human NuA4/TIP60 histone acetyltransferase (HAT) complex. Therefore, it is involved in expressional control by modifying accessibility of the DNA to the transcriptional machinery.^34^ It also plays a role in DNA damage repair: Upon DNA double-strand breaks, the TIP60/p400 complex is rapidly recruited to the damage site to remodel the surrounding chromatin, allowing repair enzymes to access and fix the DNA.^35^ *EP400* is highly expressed in neuronal tissues, particularly in the cerebellum, and during embryonic development. It ensures oligodendrocyte survival and is essential for myelination in the central nervous system.^36^

Along this line, biallelic pathogenic sequence variants, mostly missense changes, in *EP400* have been reported in 12 individuals with neurodevelopmental phenotypes, and five presented with developmental and epileptic encephalopathy.^37,38^ Notably, several genes in which a RE causes a movement disorder have also been linked to neurodevelopmental disorders through other types of variants. In *FGF14*, the intronic GAA RE causes SCA27B, whereas heterozygous point and loss-of-function variants cause SCA27A, a markedly earlier-onset disorder that includes developmental delay and intellectual disability.^39–42^ A comparable constellation exists for *HTT*, where the CAG expansion causes HD and biallelic loss-of-function variants have been described in individuals with a neurodevelopmental disorder.^43^ Thus, our findings add *EP400* to this group of genes in which a RE causes an adult-onset movement disorder, while other variant types cause a childhood-onset neurodevelopmental disorder.

From a methodological point of view, our study highlights the utility of STRling in the discovery of novel RE loci.^15^ To our knowledge, the pathogenic RE in *EP400* represents the first novel disease-associated RE identified using STRling. This finding demonstrates that STRling can detect candidate disease loci from short-read genome sequencing data alone, without relying on a curated catalog of known repeat loci. A limitation of this approach is that STRling detects repeat motifs of up to six base pairs only. However, only a small minority of the RE disorders described to date in STRipy are caused by motifs exceeding this length. It is likely that additional RE disorders remain to be identified by repeat callers such as STRling or ExpansionHunter Denovo, which likewise detects expansions at novel STR loci that are not part of a predefined catalog.^44^

Several limitations should be considered when interpreting these findings. Larger-scale screening was restricted to individuals of European ancestry, and the frequency and relevance of *EP400* expansions in other ethnicities needs to be investigated in subsequent studies. Likewise, the “gray zone” (currently, 33–36 repeat units, i.e., repeat numbers neither found in controls nor in patients) needs to be defined in larger, ethnically more diverse cohorts. Further, all of the individuals in our multiplex families are members of the same generation and intergenerational stability could not be addressed, which needs to be done when respective samples become available.

Samples investigated in this study were biobanked many years ago, and clinical information is limited. Detailed follow-up studies and reverse phenotyping of expansion carriers are warranted to fully elucidate the associated phenotype, including potential red flags and co-occurring other phenotypes. Since EP400 has also been linked to DNA damage repair, it is conceivable that it also influences the risk of cancer. Furthermore, it will be interesting to investigate the molecular consequences of the RE on the expression of *EP400* itself, but also of target genes that are regulated by EP400’s histone modifications. Polyglutamine diseases do not share a single pathological mechanism, and the pathways involved are complex. Protein-mediated toxicity and altered RNA metabolism are two examples among several proposed mechanisms, with downstream consequences including transcriptional dysregulation, impaired protein clearance, disturbed energy homeostasis and defective axonal transport.^45–47^ We did not address these mechanisms in this study, and the pathological mechanism of the *EP400* expansion therefore remains to be determined.

Taken together, we add *EP400*-related ataxia to the group of polyglutamine RE disorders. In this study, we combined a genome-wide, hypothesis-free discovery framework for REs using short-read genome data, independent clinical replication, family-based segregation evidence and long-read resolution of the repeat structure to identify and characterize this novel CAG RE disorder that causes an autosomal dominant form of SCA. This has immediate diagnostic implications for patients with unsolved dominant ataxia. Future studies will be needed to define the global distribution of *EP400*-related ataxia. We are still in exciting times, where sequencing and bioinformatic innovations drive the discovery of novel disease causes, helping us improve genetic testing by increasing the diagnostic yield. The required next step is to understand the underlying disease mechanism as a basis for mechanism-specific therapeutic interventions.

## Data availability

Raw data were generated at the Competence Centre for Genomic Analysis in Kiel, Germany, and are available from the corresponding author on request.

## Acknowledgments

We thank all patients and their families for participating in this study. During the preparation of this work, the authors used Claude (Opus 5, Anthropic) to improve the readability and language of the manuscript and BioRender AI (BioRender) to create a draft of the thumbnail image. After using these tools, the authors reviewed and edited all content as needed and take full responsibility for the content of the published article.

## Supplementary material

Supplementary material is available at *Brain* online.

## References

1. Chen Z, Morris HR, Polke J, et al. Repeat expansion disorders. Pract Neurol. 2025;25:204-216. doi:10.1136/pn-2023-003938

2. Depienne C, Mandel JL. 30 years of repeat expansion disorders: what have we learned and what are the remaining challenges? Am J Hum Genet. 2021;108:764–785. doi:10.1016/j.ajhg.2021.03.011

3. Coarelli G, Coutelier M, Durr A. Autosomal dominant cerebellar ataxias: new genes and progress towards treatments. Lancet Neurol. 2023;22:735–749. doi:10.1016/S1474-4422(23)00068-6

4. Halman A, Dolzhenko E, Oshlack A. STRipy: a graphical application for enhanced genotyping of pathogenic short tandem repeats in sequencing data. Hum Mutat. 2022;43:859–868. doi:10.1002/humu.24382

5. Perlman S. Hereditary ataxia overview. In: Adam MP, Bick S, Mirzaa GM, et al., eds. GeneReviews. University of Washington, Seattle; 1993-2026. Accessed 1 October 2026. https://www.ncbi.nlm.nih.gov/books/NBK1138/

6. Sullivan R, Yau WY, O’Connor E, Houlden H. Spinocerebellar ataxia: an update. J Neurol. 2019;266:533–544. doi:10.1007/s00415-018-9076-4

7. Paulson HL, Shakkottai VG, Clark HB, Orr HT. Polyglutamine spinocerebellar ataxias – from genes to potential treatments. Nat Rev Neurosci. 2017;18:613–626. doi:10.1038/nrn.2017.92

8. Pellerin D, Iruzubieta P, Xu IRL, et al. Recent advances in the genetics of ataxias: an update on novel autosomal dominant repeat expansions. Curr Neurol Neurosci Rep. 2025;25:16. doi:10.1007/s11910-024-01400-8

9. Tenorio RB, Camargo CHF, Donis KC, Almeida CCB, Teive HAG. Diagnostic yield of NGS tests for hereditary ataxia: a systematic review. Cerebellum. 2024;23:1552–1565. doi:10.1007/s12311-023-01629-y

10. Akçimen F, Daida K, Lange LM, et al. Long-read sequencing identifies *FGF14* repeat expansions in Parkinson’s disease. Brain. 2026;149:1514–1521. doi:10.1093/brain/awaf456

11. Alvarez Jerez P, Daida K, Miano-Burkhardt A, et al. Profiling complex repeat expansions in *RFC1* in Parkinson’s disease. NPJ Parkinsons Dis. 2024;10:108. doi:10.1038/s41531-024-00723-0

12. Shi CH, Fan Y, Yang J, et al. *NOTCH2NLC* intermediate-length repeat expansions are associated with Parkinson disease. Ann Neurol. 2021;89:182–187. doi:10.1002/ana.25925

13. Wang L, Milton M, Fearnley LG, Bhalala OG, Bahlo M, Rafehi H. Identification of expanded and interrupted *ATXN2* repeat expansions in Parkinson’s disease and Lewy body dementia cohorts. NPJ Parkinsons Dis. 2025;11:341. doi:10.1038/s41531-025-01188-5

14. Thomsen M, Borsche M, Yépez VA, et al. Multi-omics characterization of a *KIF1C* structural variant in a patient with a complex movement disorder partially responsive to deep brain stimulation. Cerebellum. 2026;25:41. doi:10.1007/s12311-026-01963-x

15. Dashnow H, Pedersen BS, Hiatt L, et al. STRling: a k-mer counting approach that detects short tandem repeat expansions at known and novel loci. Genome Biol. 2022;23:257. doi:10.1186/s13059-022-02826-4

16. Benjamini Y, Hochberg Y. Controlling the false discovery rate: a practical and powerful approach to multiple testing. J R Stat Soc Series B Stat Methodol. 1995;57:289–300. doi:10.1111/j.2517-6161.1995.tb02031.x

17. Weisburd B, Dolzhenko E, Bennett MF, et al. Defining a tandem repeat catalog and variation clusters for genome-wide analyses and population databases. Am J Hum Genet. 2026;113:915–928. doi:10.1016/j.ajhg.2026.03.020

18. Fazal S, Danzi MC, Xu I, et al. RExPRT: a machine learning tool to predict pathogenicity of tandem repeat loci. Genome Biol. 2024;25:39. doi:10.1186/s13059-024-03171-4

19. Robinson JT, Thorvaldsdóttir H, Winckler W, et al. Integrative genomics viewer. Nat Biotechnol. 2011;29:24-26. doi:10.1038/nbt.1754

20. Kasten M, Hagenah J, Graf J, et al. Cohort profile: a population-based cohort to study non-motor symptoms in parkinsonism (EPIPARK). Int J Epidemiol. 2013;42:128–128k. doi:10.1093/ije/dys202

21. Menon RP, Nethisinghe S, Faggiano S, et al. The role of interruptions in polyQ in the pathology of SCA1. PLoS Genet. 2013;9:e1003648. doi:10.1371/journal.pgen.1003648

22. De Coster W, Höijer I, Bruggeman I, et al. Visualization and analysis of medically relevant tandem repeats in nanopore sequencing of control cohorts with pathSTR. Genome Res. 2024;34:2074–2080. doi:10.1101/gr.279265.124

23. Dolzhenko E, Deshpande V, Schlesinger F, et al. ExpansionHunter: a sequence-graph-based tool to analyze variation in short tandem repeat regions. Bioinformatics. 2019;35:4754–4756. doi:10.1093/bioinformatics/btz431

24. Milovanović A, Dragašević-Mišković N, Thomsen M, et al. *RFC1* and *FGF14* repeat expansions in Serbian patients with cerebellar ataxia. Mov Disord Clin Pract. 2024;11:626–633. doi:10.1002/mdc3.14020

25. Chung MY, Ranum LP, Duvick LA, Servadio A, Zoghbi HY, Orr HT. Evidence for a mechanism predisposing to intergenerational CAG repeat instability in spinocerebellar ataxia type I. Nat Genet. 1993;5:254–258. doi:10.1038/ng1193-254

26. Sobczak K, Krzyzosiak WJ. CAG repeats containing CAA interruptions form branched hairpin structures in spinocerebellar ataxia type 2 transcripts. J Biol Chem. 2005;280:3898–3910. doi:10.1074/jbc.M409984200

27. Wright GEB, Collins JA, Kay C, et al. Length of uninterrupted CAG, independent of polyglutamine size, results in increased somatic instability, hastening onset of Huntington disease. Am J Hum Genet. 2019;104:1116–1126. doi:10.1016/j.ajhg.2019.04.007

28. Danzi MC, Xu IRL, Fazal S, et al. Population-scale variability at short tandem repeat loci reveals pathogenicity signature. bioRxiv. [Preprint] doi:10.1101/2025.01.06.631535

29. Dalski A, Pauly MG, Hanssen H, et al. Repeat length in spinocerebellar ataxia type 4 (SCA4) predicts age at onset and disease severity. J Neurol. 2024;271:6289–6300. doi:10.1007/s00415-024-12600-0

30. Hoffman-Zacharska D, Sulek A. The new face of dynamic mutation – the CAA [CAG]n CAA CAG motif as a mutable unit in the *TBP* gene causative for spino-cerebellar ataxia type 17. Int J Mol Sci. 2024;25:8190. doi:10.3390/ijms25158190

31. Yu Z, Zhu Y, Chen-Plotkin AS, et al. PolyQ repeat expansions in *ATXN2* associated with ALS are CAA interrupted repeats. PLoS One. 2011;6:e17951. doi:10.1371/journal.pone.0017951

32. Nethisinghe S, Garcia-Moreno H, Alwan J, Labrum R, Giunti P. Role of repeat tract structure and the rs7158733 SNP in spinocerebellar ataxia 3. Int J Mol Sci. 2025;26:9836. doi:10.3390/ijms26209836

33. Li LB, Yu Z, Teng X, Bonini NM. RNA toxicity is a component of ataxin-3 degeneration in *Drosophila*. Nature. 2008;453:1107–1111. doi:10.1038/nature06909

34. Yang Z, Mameri A, Cattoglio C, et al. Structural insights into the human NuA4/TIP60 acetyltransferase and chromatin remodeling complex. Science. 2024;385:eadl5816. doi:10.1126/science.adl5816

35. Pollina EA, Gilliam DT, Landau AT, et al. A NPAS4-NuA4 complex couples synaptic activity to DNA repair. Nature. 2023;614:732–741. doi:10.1038/s41586-023-05711-7

36. Elsesser O, Fröb F, Küspert M, et al. Chromatin remodeler *Ep400* ensures oligodendrocyte survival and is required for myelination in the vertebrate central nervous system. Nucleic Acids Res. 2019;47:6208–6224. doi:10.1093/nar/gkz376

37. Luo S, Wang PY, Zhou P, et al. Variants in *EP400*, encoding a chromatin remodeler, cause epilepsy with neurodevelopmental disorders. Am J Hum Genet. 2025;112:87–105. doi:10.1016/j.ajhg.2024.11.010

38. Liang XY, Meng XH, Wu WC, et al. De novo *SRCAP* variants cause developmental and epileptic encephalopathy and the phenotypic spectrum. Epilepsia. 2026;67:846–861. doi:10.1111/epi.18695

39. Rafehi H, Read J, Szmulewicz DJ, et al. An intronic GAA repeat expansion in *FGF14* causes the autosomal-dominant adult-onset ataxia SCA50/ATX-FGF14. Am J Hum Genet. 2023;110:105–119. doi:10.1016/j.ajhg.2022.11.015

40. van Swieten JC, Brusse E, de Graaf BM, et al. A mutation in the fibroblast growth factor 14 gene is associated with autosomal dominant cerebellar ataxia. Am J Hum Genet. 2003;72:191–199. doi:10.1086/345488

41. Paucar M, Lundin J, Alshammari T, et al. Broader phenotypic traits and widespread brain hypometabolism in spinocerebellar ataxia 27. J Intern Med. 2020;288:103–115. doi:10.1111/joim.13052

42. Ceroni F, Osborne D, Clokie S, et al. Analysis of fibroblast growth factor 14 (*FGF14*) structural variants reveals the genetic basis of the early onset nystagmus locus NYS4 and variable ataxia. Eur J Hum Genet. 2023;31:353–359. doi:10.1038/s41431-022-01197-5

43. Jung R, Lee Y, Barker D, et al. Mutations causing Lopes-Maciel-Rodan syndrome are huntingtin hypomorphs. Hum Mol Genet. 2021;30:135–148. doi:10.1093/hmg/ddaa283

44. Dolzhenko E, Bennett MF, Richmond PA, et al. ExpansionHunter Denovo: a computational method for locating known and novel repeat expansions in short-read sequencing data. Genome Biol. 2020;21:102. doi:10.1186/s13059-020-02017-z

45. Villavicencio Gonzalez E, Zoghbi HY. Pathogenesis of polyglutamine diseases: piecing together a complex molecular puzzle. J Exp Med. 2026;223:e20241336. doi:10.1084/jem.20241336

46. Nóbrega C, Marcelo A, Rajado AT, et al. Molecular hallmarks of neurodegeneration in polyglutamine spinocerebellar ataxias. Cell Death Dis. 2025;16:826. doi:10.1038/s41419-025-08154-2

47. Malik I, Kelley CP, Wang ET, Todd PK. Molecular mechanisms underlying nucleotide repeat expansion disorders. Nat Rev Mol Cell Biol. 2021;22:589–607. doi:10.1038/s41580-021-00382-6

