## Supplementary material for "A coding CAG repeat expansion in *EP400* causes Spinocerebellar Ataxia": Supplmenentary material

### **PCR-Protocol 1: M13-tailed PCR for screening (Taq)**

*Taq*-based PCR used for screening of the repeat locus by fragment analysis. The forward primer carries a 5′ M13F tail, shown in gray; the gene-specific part follows in black. The fluorescent label is carried by the universal M13F primer.

#### **Primers**

| **Primer** |  |
| --- | --- |
| EP400-M13F | CACGACGTTGTAAAACGACATCTGAAGGTGGGCGAGTATC |
| EP400-R | ATGGTCCTCTGGGAAATGC |

#### **Reaction setup**

| **Component** | **Volume (µL)** |
| --- | --- |
| 10× Incubation Mix with MgCl2 | 1.5 |
| dNTP mix | 3.0 |
| Primer M13-F | 0.4 |
| Primer R | 0.4 |
| Betaine | 1.0 |
| DMSO | 1.0 |
| M13 | 0.3 |
| Taq DNA polymerase (5 U/µL) | 0.1 |
| H2O | 5.8 |
| Genomic DNA (100 ng/µL) | 1.5 |
| Total reaction volume | 15.0 |

#### **Thermal cycling**

The ramp rate of the thermal cycler was set to 6 °C/s. Annealing was performed as a two-step touchdown (64 °C, 60 °C).

| **Step** | **Temperature** | **Time** | **Cycles** |
| --- | --- | --- | --- |
| Initial denaturation | 95 °C | 5 min | 1 |
| Denaturation  Annealing  Extension | 95 °C  64 °C  72 °C | 1 min  30 s  3 min | 10× |
| Denaturation  Annealing  Extension | 95 °C  60 °C  72 °C | 1 min  30 s  3 min | 25× |
| Final extension | 72 °C | 10 min | 1 |
| Hold | 4 °C | ∞ | – |

### **PCR-Protocol 2: High-fidelity PCR for long-read sequencing (Phusion)**

Phusion based high-fidelity PCR used to generate the amplicon for long-read sequencing. Both primers are gene-specific and carry no tail.

#### **Primers**

| **Primer** |  |
| --- | --- |
| EP400-F | ATCTGAAGGTGGGCGAGTATC |
| EP400-R | ATGGTCCTCTGGGAAATGC |

#### **Reaction setup**

| **Component** | **Volume (µL)** |
| --- | --- |
| Phusion Flash High-Fidelity PCR Master Mix | 12.50 |
| Primer F | 1.50 |
| Primer R | 1.50 |
| DMSO | 1.25 |
| H2O | 5.25 |
| Genomic DNA (50 ng/µL) | 3.00 |
| Total reaction volume | 25.00 |

#### **Thermal cycling**

The ramp rate of the thermal cycler was set to 3 °C/s. Annealing was performed as a three step touchdown (66 °C, 64 °C, 62 °C).

| **Step** | **Temperature** | **Time** | **Cycles** |
| --- | --- | --- | --- |
| Initial denaturation | 98 °C | 1 min | 1 |
| Denaturation  Annealing  Extension | 98 °C  66 °C  72 °C | 5 s  10 s  30 s | 10× |
| Denaturation  Annealing  Extension | 98 °C  64 °C  72 °C | 5 s  10 s  30 s | 10× |
| Denaturation  Annealing  Extension | 98 °C  62 °C  72 °C | 5 s  10 s  30 s | 15× |
| Final extension | 72 °C | 1 min | 1 |
| Hold | 4 °C | ∞ | – |

**STR marker primers**

Primer sequences of the custom microsatellite marker used for haplotype analysis with the universal M13 tail in gray.

| **Marker** | **Forward primer** | **Reverse primer** |
| --- | --- | --- |
| Custom marker | CACGACGTTGTAAAACGACTCCATCAGACGCTTGTCCAC | GGGTGAATAAATGGCACGGC |

**Table S1. Longest observed allele per repeat locus in unaffected individuals, ataxia and dystonia patients.**

Values are estimated repeat unit counts of the longer allele (allele 2); for each locus and group the maximum across all individuals is shown. Genomic coordinates refer to GRCh38. A dash indicates that no carrier was observed in the respective group.

| Chromosome | Gene | Locus | Motif | Unaffected | Ataxia | Dystonia |
| --- | --- | --- | --- | --- | --- | --- |
| chr1 | *TNFRSF8* | chr1:12124829-12124881 | AAAAC | 13.2 | – | 15.5 |
| chr1 | *FBXO42* | chr1:16350757-16350796 | AAAG | – | – | 25.9 |
| chr1 | *ALPL* | chr1:21527015-21527046 | CTTTT | 14.8 | 27.4 | 32.9 |
| chr1 | *LDLRAP1* | chr1:25589304-25589331 | AAAG | 37.2 | 30.2 | 38.4 |
| chr1 | *FOXJ3* | chr1:42221980-42222021 | AAG | – | – | 59.7 |
| chr1 | *FOXJ3* | chr1:42222034-42222035 | AAG | – | – | 35.9 |
| chr1 | *AGBL4* | chr1:49302610-49302611 | ATTTT | – | – | 14.6 |
| chr1 | *MROH7* | chr1:54699094-54699095 | CTTT | 21.0 | 22.6 | 23.9 |
| chr1 | *NFIA* | chr1:61025464-61025465 | AAAG | – | 14.1 | 11.6 |
| chr1 | *KANK4* | chr1:62278381-62278382 | CTTT | – | – | 12.9 |
| chr1 | *LEPR* | chr1:65488146-65488147 | CTTT | 20.3 | 23.7 | 21.0 |
| chr1 | *SGIP1* | chr1:66630802-66630905 | AAAG | – | 11.9 | 21.1 |
| chr1 | *LRRC7* | chr1:69781736-69781763 | AAAG | 23.0 | 14.8 | 27.7 |
| chr1 | *CASQ2* | chr1:115761402-115761403 | AAG | 38.7 | 55.5 | 45.3 |
| chr1 | *CFH* | chr1:196692715-196692716 | CTTT | – | – | 16.9 |
| chr1 | *MAP3K21* | chr1:233339277-233339278 | CTT | 18.2 | – | 20.9 |
| chr1 | *SLC35F3* | chr1:234139951-234140002 | AAAAT | 38.7 | 13.3 | 41.0 |
| chr1 | *RYR2* | chr1:237061211-237061212 | ATCT | – | – | 15.1 |
| chr1 | *RYR2* | chr1:237237967-237237987 | CCTTT | – | – | 15.3 |
| chr1 | *PLD5* | chr1:242194641-242194642 | ATCT | – | – | 21.8 |
| chr2 | *PDIA6* | chr2:10806648-10806649 | AAGG | – | 16.6 | 19.0 |
| chr2 | *VSNL1* | chr2:17622506-17622507 | AAAG | 22.0 | – | 29.1 |
| chr2 | *LBH* | chr2:30319597-30319598 | ATGG | 16.7 | 19.8 | 17.6 |
| chr2 | *PRKCE* | chr2:45744485-45744486 | CTTT | 13.9 | – | 19.0 |
| chr2 | *CRIPT* | chr2:46660706-46660707 | CTTT | 23.0 | – | 33.1 |
| chr2 | *STON1* | chr2:48564479-48564480 | CCTTCT | 9.1 | – | 17.6 |
| chr2 | *STON1-GTF2A1L* | chr2:48564479-48564480 | CCTTCT | 9.1 | – | 17.6 |
| chr2 | *ACYP2* | chr2:54197999-54198000 | ATTGT | 10.9 | 20.9 | 44.3 |
| chr2 | *EML6* | chr2:54907972-54907973 | AGAT | 12.2 | – | 18.2 |
| chr2 | *MTHFD2* | chr2:74203843-74203928 | AGTTT | 14.2 | 23.0 | 32.3 |
| chr2 | *SH3RF3* | chr2:109199956-109199957 | AATGG | – | 9.4 | 14.2 |
| chr2 | *MGAT5* | chr2:134381330-134381331 | AGAT | 19.4 | 13.4 | 28.9 |
| chr2 | *RAB3GAP1* | chr2:135117409-135117410 | CTT | – | – | 19.8 |
| chr2 | *ZEB2* | chr2:144375869-144375961 | AAAG | 15.6 | – | 18.2 |
| chr2 | *TMEFF2* | chr2:191964323-191964324 | CTTT | 32.9 | – | 40.0 |
| chr2 | *TMEFF2* | chr2:192037273-192037336 | AAAG | 38.8 | 23.9 | 52.3 |
| chr2 | *PLCL1* | chr2:198422739-198422807 | ATTTT | 17.6 | 12.3 | 39.2 |
| chr2 | *EPHA4* | chr2:221566956-221567011 | AAG | 40.2 | 43.7 | 47.4 |
| chr2 | *AGAP1* | chr2:235573061-235573062 | CTT | 55.2 | 33.8 | 72.1 |
| chr3 | *RFT1* | chr3:53065210-53065211 | CTTT | – | – | 16.0 |
| chr3 | *CACNA2D3* | chr3:54822730-54822731 | CTTT | 18.4 | 19.5 | 25.9 |
| chr3 | *TAFA1* | chr3:68120249-68120264 | CTTT | – | – | 13.8 |
| chr3 | *PPP4R2* | chr3:73018413-73018414 | ATGTT | 14.1 | 14.7 | 17.7 |
| chr3 | *CADM2* | chr3:85568490-85568491 | CTTT | 16.7 | 13.5 | 20.1 |
| chr3 | *EPHA6* | chr3:97324405-97324472 | CTTT | 13.7 | 16.2 | 17.1 |
| chr3 | *DCBLD2* | chr3:98870737-98870813 | AAAG | – | 11.5 | 13.5 |
| chr3 | *DRD3* | chr3:114156840-114156841 | CTTT | – | 15.4 | 15.0 |
| chr3 | *LSAMP* | chr3:116684503-116684504 | ATTTT | – | – | 33.3 |
| chr3 | *IQCB1* | chr3:121786170-121786211 | AAGAG | – | – | 21.9 |
| chr3 | *IQCB1* | chr3:121786170-121786215 | AAAAG | 27.6 | 35.6 | 42.8 |
| chr3 | *IQCB1* | chr3:121786191-121786193 | AAAGG | – | 22.7 | 24.1 |
| chr3 | *PARP9* | chr3:122539507-122539570 | CTTT | 23.4 | 14.2 | 25.1 |
| chr3 | *KALRN* | chr3:124286065-124286066 | CTTT | – | – | 18.6 |
| chr3 | *NEK11* | chr3:131115902-131115983 | CTTT | – | – | 12.5 |
| chr3 | *CLSTN2* | chr3:140251638-140251639 | CCTTT | – | – | 23.2 |
| chr3 | *SPTSSB* | chr3:161369339-161369340 | CTTT | 15.8 | 13.3 | 21.4 |
| chr3 | *MECOM* | chr3:169378457-169378505 | AAAG | 15.3 | 13.8 | 21.6 |
| chr3 | *FGF12* | chr3:192378026-192378120 | CTTT | – | – | 15.1 |
| chr4 | *NELFA* | chr4:2020799-2020800 | CTTT | 13.8 | – | 16.2 |
| chr4 | *RBPJ* | chr4:26210650-26210810 | CTTT | – | 11.9 | 12.5 |
| chr4 | *ARAP2* | chr4:36014309-36014310 | AAAG | – | 13.5 | 23.9 |
| chr4 | *APBB2* | chr4:41159943-41160037 | AAG | – | 62.6 | 62.5 |
| chr4 | *ATP8A1* | chr4:42555214-42555215 | ATCT | – | – | 13.8 |
| chr4 | *ADAMTS3* | chr4:72549962-72549963 | AAG | 56.2 | 105.2 | 95.9 |
| chr4 | *SHROOM3* | chr4:76608682-76608683 | ACAGC | – | – | 17.3 |
| chr4 | *LIN54* | chr4:83001858-83001859 | AAAGG | 12.7 | 18.6 | 29.4 |
| chr4 | *LIN54* | chr4:83001955-83001956 | AAAGG | 11.8 | 23.8 | 20.0 |
| chr4 | *ELF2* | chr4:139092366-139092367 | AACAT | 10.9 | 40.3 | 39.2 |
| chr4 | *NAA15* | chr4:139315006-139315055 | AGGTT | – | – | 24.8 |
| chr4 | *SETD7* | chr4:139572196-139572218 | CTT | 68.0 | 92.0 | 85.2 |
| chr4 | *SCOC* | chr4:140360993-140360994 | CTT | 26.5 | – | 38.6 |
| chr4 | *RNF150* | chr4:141000002-141000003 | AAGGAG | – | – | 12.6 |
| chr4 | *TRIM2* | chr4:153244291-153244437 | CTT | 48.3 | 22.6 | 52.6 |
| chr4 | *FSTL5* | chr4:161783707-161783708 | CTTT | 11.7 | – | 18.6 |
| chr4 | *DDX60* | chr4:168297350-168297401 | AAAG | 14.3 | 33.7 | 24.0 |
| chr5 | *SEMA5A* | chr5:9426432-9426479 | AAAAT | 24.9 | 14.9 | 30.2 |
| chr5 | *SUB1* | chr5:32568273-32568274 | AAAG | – | – | 12.7 |
| chr5 | *GHR* | chr5:42579074-42579075 | AGAT | 15.2 | 24.0 | 30.7 |
| chr5 | *GZMA* | chr5:55105972-55105973 | AAAAT | 23.7 | 33.8 | 34.5 |
| chr5 | *IPO11* | chr5:62598431-62598433 | CTTT | 15.3 | 19.9 | 28.3 |
| chr5 | *IPO11* | chr5:62598492-62598493 | CTTT | 22.2 | 23.0 | 36.2 |
| chr5 | *IQGAP2* | chr5:76496762-76496763 | CTTT | 14.4 | 13.0 | 21.4 |
| chr5 | *DMGDH* | chr5:79180630-79180631 | AAGGC | 22.8 | – | 28.4 |
| chr5 | *SPOCK1* | chr5:137518584-137518585 | AAAG | – | – | 14.3 |
| chr5 | *CTNNA1* | chr5:138740884-138740885 | CTTT | – | 13.6 | 18.5 |
| chr5 | *ECSCR* | chr5:139446465-139446558 | CTTT | 13.2 | 14.4 | 21.2 |
| chr5 | *TAF7* | chr5:141275157-141275158 | AAAG | – | – | 18.8 |
| chr5 | *JAKMIP2* | chr5:147702557-147702558 | AAAG | – | – | 18.5 |
| chr5 | *PPARGC1B* | chr5:149809226-149809392 | AGAT | 12.7 | 15.8 | 15.8 |
| chr5 | *GLRA1* | chr5:151849391-151849458 | CCTTT | – | – | 10.0 |
| chr5 | *TENM2* | chr5:167872509-167872510 | AAAG | 17.7 | – | 20.5 |
| chr5 | *WWC1* | chr5:168403006-168403007 | CTTT | 12.4 | – | 18.7 |
| chr5 | *KCNIP1* | chr5:170367417-170367418 | AAAG | 14.5 | – | 24.1 |
| chr5 | *KCNMB1* | chr5:170367417-170367418 | AAAG | 14.5 | – | 24.1 |
| chr5 | *STK10* | chr5:172056970-172056971 | AAAG | 17.4 | 17.7 | 25.8 |
| chr5 | *SH3PXD2B* | chr5:172348694-172348695 | ATCT | – | – | 14.1 |
| chr6 | *GFOD1* | chr6:13409166-13409183 | AAAG | – | – | 22.8 |
| chr6 | *CD83* | chr6:14151491-14151559 | CCTT | – | 13.6 | 16.2 |
| chr6 | *CDKAL1* | chr6:20756862-20756863 | CCTT | 13.3 | 15.1 | 20.4 |
| chr6 | *CARMIL1* | chr6:25459210-25459269 | CTTT | 44.0 | 52.8 | 74.2 |
| chr6 | *ZFAND3* | chr6:38041806-38041807 | CCTTCT | – | – | 9.8 |
| chr6 | *TRERF1* | chr6:42277926-42277927 | AAG | 15.7 | – | 28.0 |
| chr6 | *TENT5A* | chr6:81680201-81680202 | AAAG | 11.8 | – | 20.1 |
| chr6 | *FRK* | chr6:115958718-115958719 | AAAG | 18.7 | 16.5 | 21.8 |
| chr6 | *HDDC2* | chr6:125260982-125260983 | AAAG | – | – | 23.6 |
| chr6 | *TPD52L1* | chr6:125260982-125260983 | AAAG | – | – | 23.6 |
| chr6 | *SLC18B1* | chr6:132792291-132792304 | AAAG | – | – | 17.4 |
| chr6 | *MAP3K5* | chr6:136585476-136585489 | CTTT | – | – | 15.6 |
| chr6 | *SASH1* | chr6:148387590-148387591 | CTTT | 30.2 | 23.6 | 31.9 |
| chr6 | *TAB2* | chr6:149253995-149253996 | AAAG | 13.4 | 13.2 | 13.2 |
| chr6 | *MTHFD1L* | chr6:150994077-150994097 | AAAG | 12.7 | – | 15.4 |
| chr6 | *SYNE1* | chr6:152139707-152139752 | AAAG | 14.4 | 16.1 | 17.3 |
| chr6 | *IGF2R* | chr6:159980183-159980239 | AAAG | – | – | 55.3 |
| chr6 | *PACRG* | chr6:163269977-163269978 | AAAG | 24.9 | 18.6 | 27.4 |
| chr7 | *GNA12* | chr7:2812636-2812697 | ACATC | 22.9 | 13.3 | 22.9 |
| chr7 | *DGKB* | chr7:14170145-14170204 | AAAG | 27.1 | – | 41.3 |
| chr7 | *PDE1C* | chr7:32147237-32147331 | AAAG | – | 17.5 | 20.6 |
| chr7 | *TNS3* | chr7:47389076-47389077 | AAG | 26.7 | 46.9 | 71.5 |
| chr7 | *GALNT17* | chr7:71592563-71592564 | AAAAT | – | – | 13.3 |
| chr7 | *HIP1* | chr7:75595187-75595254 | CTTT | – | 13.4 | 23.0 |
| chr7 | *ZNF804B* | chr7:88854396-88854397 | CCTT | – | – | 19.1 |
| chr7 | *ZNF804B* | chr7:89171288-89171289 | CTT | 17.9 | – | 21.1 |
| chr7 | *MET* | chr7:116755004-116755005 | AAAG | 13.7 | 14.5 | 18.0 |
| chr7 | *CPED1* | chr7:121047646-121047717 | CTT | – | 20.3 | 16.2 |
| chr7 | *PLXNA4* | chr7:132132380-132132381 | CTGTT | 33.4 | 13.7 | 38.3 |
| chr7 | *PLXNA4* | chr7:132188951-132188995 | AAAGG | 15.1 | 11.9 | 22.4 |
| chr7 | *DGKI* | chr7:137767485-137767486 | AAGAG | – | – | 11.2 |
| chr7 | *MGAM* | chr7:141913869-141913870 | AAAAT | 12.3 | 10.9 | 18.0 |
| chr8 | *CSGALNACT1* | chr8:19599478-19599479 | AAAG | – | – | 15.5 |
| chr8 | *CSGALNACT1* | chr8:19666943-19667019 | AAAG | 14.6 | – | 16.9 |
| chr8 | *EPHX2* | chr8:27501324-27501395 | CTT | 35.2 | 28.9 | 64.7 |
| chr8 | *CHRNA6* | chr8:42766494-42766521 | AAAG | – | – | 24.9 |
| chr8 | *MTERF3* | chr8:96250615-96250683 | AAG | 48.1 | 16.2 | 74.0 |
| chr8 | *MTERF3* | chr8:96250674-96250675 | AAGGAG | 11.3 | – | 11.7 |
| chr8 | *ANGPT1* | chr8:107370218-107370219 | AAAG | 19.2 | 16.6 | 22.3 |
| chr8 | *CSMD3* | chr8:112370045-112370084 | AAG | – | 17.5 | 22.2 |
| chr9 | *PTPRD* | chr9:10060897-10060898 | CTTT | – | – | 12.6 |
| chr9 | *CCDC171* | chr9:15883018-15883019 | CCTT | 11.6 | – | 12.8 |
| chr9 | *SLC24A2* | chr9:19636315-19636316 | CTTT | – | 15.2 | 20.9 |
| chr9 | *MLLT3* | chr9:20414279-20414402 | CTG | – | 20.0 | – |
| chr9 | *APBA1* | chr9:69658215-69658283 | CTTT | – | – | 16.4 |
| chr9 | *DAPK1* | chr9:87575263-87575274 | AAAAT | – | 9.7 | 21.7 |
| chr9 | *DAB2IP* | chr9:121642029-121642030 | CTTT | 12.1 | – | 17.4 |
| chr9 | *RABGAP1* | chr9:123077610-123077693 | ATTGT | 30.1 | – | 31.3 |
| chr9 | *NCS1* | chr9:130176138-130176197 | CTTT | – | – | 34.1 |
| chr10 | *SFMBT2* | chr10:7315037-7315038 | AAAG | 12.5 | – | 14.9 |
| chr10 | *ITIH5* | chr10:7566826-7566827 | AAG | – | – | 34.1 |
| chr10 | *CELF2* | chr10:11197012-11197013 | AAAG | 16.9 | 14.0 | 27.9 |
| chr10 | *CAMK1D* | chr10:12801370-12801394 | ATCT | 15.7 | 11.6 | 17.4 |
| chr10 | *PLXDC2* | chr10:20072459-20072460 | AAAG | – | 12.5 | 13.4 |
| chr10 | *MSRB2* | chr10:23134542-23134611 | AAAG | – | – | 15.5 |
| chr10 | *ENKUR* | chr10:25054457-25054458 | CTTT | – | 13.6 | 11.8 |
| chr10 | *ARHGAP22* | chr10:48655091-48655092 | CCTCT | 9.7 | – | 15.9 |
| chr10 | *CTNNA3* | chr10:67302018-67302019 | AAAG | – | – | 29.6 |
| chr10 | *HK1* | chr10:69332357-69332392 | CTTT | 27.2 | – | 45.8 |
| chr10 | *NRG3* | chr10:82761972-82761973 | CTTT | 18.1 | – | 21.0 |
| chr10 | *SLIT1* | chr10:97166589-97166590 | AAAG | – | – | 16.5 |
| chr10 | *HPSE2* | chr10:99132187-99132194 | AAAG | 14.3 | 20.2 | 19.6 |
| chr10 | *HPSE2* | chr10:99132241-99132242 | AAAG | 14.4 | 15.5 | 34.1 |
| chr11 | *ABCC8* | chr11:17450313-17450314 | CTTT | 11.6 | – | 16.8 |
| chr11 | *LRRC4C* | chr11:40984366-40984367 | AAAG | 11.7 | 13.9 | 17.0 |
| chr11 | *DDB1* | chr11:61311258-61311259 | AACAT | 31.5 | – | 41.1 |
| chr11 | *SHANK2* | chr11:70599896-70599897 | AAAG | 13.4 | 15.5 | 21.1 |
| chr11 | *SHANK2* | chr11:70903376-70903377 | AAAAT | 25.2 | – | 38.9 |
| chr11 | *DLG2* | chr11:84626888-84626888 | ATTTT | 11.0 | – | 27.9 |
| chr11 | *C11orf71* | chr11:114394184-114394311 | CTTTT | 15.6 | 9.9 | 27.5 |
| chr11 | *GRAMD1B* | chr11:123457111-123457139 | AAAG | 23.3 | 25.3 | 34.9 |
| chr11 | *NTM* | chr11:131789488-131789489 | AAGGAG | 11.7 | – | 14.0 |
| chr12 | *ERC1* | chr12:1121848-1121849 | ATCTCT | – | – | 118.7 |
| chr12 | *VWF* | chr12:5964219-5964220 | ACAT | – | 20.3 | 18.1 |
| chr12 | *CLSTN3* | chr12:7148156-7148215 | AAAG | – | – | 45.8 |
| chr12 | *RERG* | chr12:15161138-15161229 | AAAG | – | 15.2 | 19.1 |
| chr12 | *STAT6* | chr12:57100674-57100675 | AAAG | – | 13.0 | 12.3 |
| chr12 | *MSRB3* | chr12:65396704-65396755 | AAAG | – | – | 22.6 |
| chr12 | *GRIP1* | chr12:66414935-66414989 | AAAAT | 22.8 | – | 34.1 |
| chr12 | *NAV3* | chr12:77755511-77755512 | CCTTT | – | 11.2 | 11.6 |
| chr12 | *ACSS3* | chr12:81045057-81045058 | CTTT | 14.0 | – | 18.4 |
| chr12 | *ANKS1B* | chr12:99139381-99139382 | CTTT | – | 13.0 | 16.7 |
| chr12 | *RPH3A* | chr12:112712972-112712973 | CCTTCT | – | – | 17.3 |
| chr12 | *RPH3A* | chr12:112713102-112713155 | CTT | 89.1 | 46.7 | 98.7 |
| chr12 | *NOS1* | chr12:117398692-117398703 | AAAAT | – | 20.4 | 25.6 |
| chr12 | *KSR2* | chr12:117947206-117947259 | AAAG | 15.3 | – | 20.5 |
| chr12 | *VSIG10* | chr12:118133154-118133185 | ATTTC | 14.3 | 10.2 | 19.5 |
| chr12 | *VSIG10* | chr12:118133227-118133228 | ATTTC | 23.5 | – | 26.9 |
| chr12 | *RFLNA* | chr12:124296125-124296125 | CTTT | 17.7 | 17.4 | 25.9 |
| chr12 | *GLT1D1* | chr12:128879378-128879461 | CTTT | 14.0 | – | 25.4 |
| chr12 | *EP400* | chr12:132062523-132062524 | AGC | – | 40.2 | – |
| chr13 | *WASF3* | chr13:26559788-26559811 | CTTT | 20.4 | 17.7 | 42.1 |
| chr13 | *FLT1* | chr13:28453110-28453111 | CCTTT | – | – | 14.4 |
| chr13 | *RB1* | chr13:48459947-48459948 | CTTTT | – | – | 21.6 |
| chr13 | *DACH1* | chr13:71748923-71748924 | CTTT | 13.3 | 12.1 | 19.0 |
| chr13 | *ABCC4* | chr13:95196666-95196667 | AAGG | 13.3 | 12.3 | 16.1 |
| chr13 | *ABCC4* | chr13:95244664-95244672 | AAAG | – | 28.4 | 17.3 |
| chr13 | *TNFSF13B* | chr13:108258599-108258600 | ATATGT | – | 9.0 | 7.7 |
| chr14 | *NYNRIN* | chr14:24406201-24406202 | AAAAT | 14.0 | – | 25.9 |
| chr14 | *EGLN3* | chr14:33978246-33978247 | AAAG | 17.3 | 15.7 | 22.5 |
| chr14 | *RCOR1* | chr14:102632644-102632645 | CCTTT | – | – | 11.9 |
| chr15 | *GALK2* | chr15:49299709-49299710 | CTTT | 14.6 | – | 19.1 |
| chr15 | *TCF12* | chr15:57075835-57075863 | CTTT | 47.2 | 31.2 | 67.1 |
| chr15 | *PPCDC* | chr15:75097199-75097254 | CTTT | 23.4 | 41.9 | 44.4 |
| chr16 | *PRKCB* | chr16:24021055-24021072 | CTTT | – | 16.2 | 12.4 |
| chr16 | *HS3ST4* | chr16:25828214-25828215 | CTTT | 11.5 | 12.2 | 19.4 |
| chr16 | *GSG1L* | chr16:27888508-27888509 | CTTT | 21.3 | – | 23.1 |
| chr16 | *ATXN1L* | chr16:71832263-71832306 | AAG | 62.9 | 72.4 | 67.5 |
| chr16 | *WWOX* | chr16:78723660-78723661 | CTTTT | 11.8 | 10.5 | 14.9 |
| chr17 | *KIAA0753* | chr17:6591045-6591046 | AAG | 23.3 | 38.5 | 27.8 |
| chr17 | *TP53* | chr17:7664361-7664362 | ATCT | – | – | 15.3 |
| chr17 | *CFAP52* | chr17:9631052-9631053 | AAAG | 24.7 | – | 45.3 |
| chr17 | *GAS7* | chr17:10046876-10046877 | AAAG | – | – | 14.7 |
| chr17 | *SLFN12L* | chr17:35530441-35530442 | AAAG | 16.1 | – | 17.3 |
| chr17 | *CACNB1* | chr17:39183337-39183366 | AAG | – | 75.9 | 87.2 |
| chr17 | *CDK12* | chr17:39498900-39498901 | CCTT | – | – | 20.6 |
| chr17 | *CDK12* | chr17:39499045-39499046 | CTTT | 46.0 | 57.4 | 47.8 |
| chr17 | *ABCC3* | chr17:50673900-50673977 | CTTT | 14.3 | 13.5 | 21.4 |
| chr17 | *CA10* | chr17:51831667-51831732 | AGC | 88.0 | 89.3 | 106.5 |
| chr17 | *MSI2* | chr17:57450255-57450322 | AAAG | 14.2 | 19.8 | 34.1 |
| chr17 | *TEX2* | chr17:64219508-64219554 | AAAAT | 11.0 | – | 15.6 |
| chr17 | *SLC39A11* | chr17:73004178-73004244 | AAAG | 14.5 | 14.8 | 23.2 |
| chr17 | *CD300A* | chr17:74482697-74482733 | CTTT | 20.9 | – | 32.8 |
| chr17 | *C1QTNF1* | chr17:79030514-79030515 | CTTT | – | – | 14.7 |
| chr18 | *NDC80* | chr18:2614439-2614656 | AAAG | 32.7 | – | 46.8 |
| chr18 | *EPB41L3* | chr18:5566714-5566715 | ATTCT | 19.1 | – | 20.4 |
| chr18 | *EPB41L3* | chr18:5566727-5566728 | ATTCC | 22.8 | 22.4 | 24.2 |
| chr18 | *L3MBTL4* | chr18:5958111-5958112 | AAG | 32.7 | 24.2 | 36.1 |
| chr18 | *RBBP8* | chr18:23022603-23022671 | AAAAT | – | – | 33.9 |
| chr18 | *GAREM1* | chr18:32145692-32145727 | CTTT | 17.9 | 36.0 | 29.1 |
| chr18 | *GRP* | chr18:59226985-59227068 | CTTT | – | – | 13.3 |
| chr18 | *RNF152* | chr18:61844141-61844141 | AAAG | – | – | 12.5 |
| chr18 | *BCL2* | chr18:63169336-63169337 | CTTT | 12.0 | 19.4 | 19.0 |
| chr19 | *SBNO2* | chr19:1165099-1165100 | AGG | – | – | 26.5 |
| chr19 | *ARHGEF18* | chr19:7361993-7361994 | AAGGAG | 9.2 | – | 10.8 |
| chr19 | *WIZ* | chr19:15425913-15425914 | AGG | 20.6 | – | 23.1 |
| chr19 | *CYP4F3* | chr19:15650660-15650797 | CTTT | – | 14.6 | 12.4 |
| chr19 | *NWD1* | chr19:16737936-16738013 | AAAAG | 14.1 | 10.8 | 16.3 |
| chr19 | *COPE* | chr19:18902825-18902826 | AAAG | – | – | 26.2 |
| chr19 | *DPY19L3* | chr19:32447807-32447808 | AGAT | 16.4 | – | 23.0 |
| chr19 | *TMEM143* | chr19:48334496-48334497 | CTTT | 15.7 | – | 17.1 |
| chr19 | *SHANK1* | chr19:50707876-50707965 | CTTTT | 18.0 | 13.9 | 23.5 |
| chr20 | *PLCB1* | chr20:8629910-8629911 | CTTT | – | – | 13.4 |
| chr20 | *PCSK2* | chr20:17347919-17347920 | AAAG | 11.5 | 14.3 | 18.5 |
| chr20 | *RIN2* | chr20:19844641-19844642 | CTT | 15.5 | – | 33.3 |
| chr20 | *RIPOR3* | chr20:50666183-50666226 | CTTTT | – | – | 39.9 |
| chr20 | *RIPOR3* | chr20:50666198-50666200 | CCTTT | 18.5 | 22.0 | 19.2 |
| chr20 | *CDH4* | chr20:61714026-61714027 | ATGTT | – | – | 15.4 |
| chr20 | *CDH4* | chr20:61714058-61714059 | ATGTT | – | – | 10.6 |
| chr20 | *RTEL1* | chr20:63693519-63693520 | ACC | – | 19.7 | 22.5 |
| chr22 | *SEZ6L* | chr22:26348566-26348567 | AAAG | – | – | 14.2 |
| chr22 | *TTC28* | chr22:28537293-28537341 | AAAAT | 17.3 | 19.5 | 16.1 |
| chr22 | *TTC28* | chr22:28537331-28537332 | AACAT | – | – | 11.5 |
| chrX | *ARHGAP6* | chrX:11174634-11174635 | CTTT | – | 15.2 | 17.8 |
| chrX | *ARHGAP6* | chrX:11590791-11590792 | AAAAG | 10.3 | – | 13.7 |
| chrX | *ARHGAP6* | chrX:11590833-11590862 | AAAG | – | – | 16.6 |
| chrX | *FRMPD4* | chrX:12193835-12193836 | AAAG | 14.1 | – | 31.2 |
| chrX | *ACOT9* | chrX:23759749-23759750 | AAAG | 12.7 | 13.6 | 26.3 |
| chrX | *COL4A5* | chrX:108526595-108526596 | CTTT | – | 26.0 | 24.9 |
| chrX | *SEPTIN6* | chrX:119637640-119637641 | ATCC | – | 11.8 | 12.4 |
